# Oestradiol modulates the brain age gap

**DOI:** 10.64898/2025.12.23.25342899

**Authors:** Anna F. Denninger, Tobias Kaufmann, Inger Sundström-Poromaa, Birgit Derntl, Lydia Kogler

**Affiliations:** Department of Psychiatry and Psychotherapy, Center for Mental Health (TüCMH), University of Tübingen, Tübingen, Germany; German Center for Mental Health (DZPG), partner site Tübingen, Tübingen, Germany; Centre for precision Psychiatry, University of Oslo, Oslo, Norway; Department of Women’s and Children’s Health, Uppsala University, Uppsala, Sweden; LEAD Graduate School, University of Tübingen, Germany

**Author notes:** Corresponding authors: Anna F. Denninger & Dr. Lydia Kogler, Department of Psychiatry & Psychotherapy, Women’s Mental Health and Brain Function, Tübingen Center for Mental Health, University of Tübingen, Calwerstrasse 14, 72076 Tübingen, Germany.

**Keywords:** Oestradiol, brain age gap, menstrual cycle, emotion regulation, self-esteem, resilience

## Abstract

The brain age gap (BAG) — the difference between chronological and neuroimaging-based predicted brain age — has emerged as a sensitive biomarker of brain health. A higher BAG, reflecting an older-appearing brain, has been linked to cognitive decline, neurodegenerative disease, and mental disorders. Whether such apparent aging can be mitigated by targeted interventions remains unclear. Oestradiol (E2), a sex hormone fluctuating across the menstrual cycle and known for its neuromodulatory and cognitive effects, may represent one such target intervention.

To test this, we conducted a double-blind, randomized, placebo-controlled crossover study in 28 premenopausal females. Each participant was assessed twice during the follicular phase, with at least two cycles between sessions, and received either 12 mg E2-valerate or a placebo before undergoing a T1-weighted MRI scan. BAG was estimated using a pre-trained Simple Fully Convolutional Network model.

Predicted brain age was significantly younger following E2 administration compared to placebo. Exploratory analyses indicated that this effect may be moderated by resilience factors for mental health: Higher self-esteem and use of greater social support seeking was nominally associated with lower BAG, whereas females applying more expressive suppression showed more apparent brain aging.

These findings suggest that even short-term increases in E2, together with resilience factors, can influence brain age estimates. Thus, E2 interventions may provide a targeted approach to support both brain and mental health. These findings underscore the urgent need for further research into the impact of E2 on mental health across the female lifespan.

## Introduction

The brain age gap (BAG), defined as the difference between an individual’s chronological age and neuroimaging-based predicted brain age, has emerged as a neuro-biomarker for brain health (Kaufmann et al., 2019; Leonardsen et al., 2022; Seitz-Holland et al., 2024). A larger positive BAG, indicating an older-appearing brain, has previously been associated with neurodegenerative diseases, and mental disorders (Han et al., 2022; Karim et al., 2021; Kaufmann et al., 2019; Leonardsen et al., 2022; Seitz-Holland et al., 2024). The degree to which apparent aging-effects in the brain might be attenuated or even reverted by targeted interventions remains open. A possible treatment target might be oestradiol (E2), which has previously been implicated for its positive neuromodulatory effects and possible support of cognitive and emotional functioning (Derntl et al., 2024; Hampson & Morley, 2013; Luine, 2014; Rehbein et al., 2021).

E2 is a sex hormone that fluctuates across the female life span. In naturally cycling females E2 undergoes cyclic fluctuations and peaks during pregnancy, whereas after menopause these fluctuations cease, resulting in a stable low-hormone state (Pletzer et al. 2025, Barth et al., 2023). Naturally high-E2 phases (pre-ovulation) of the menstrual cycle have been linked to younger predicted brain age (Franke et al., 2015). However, other hormones such as progesterone and testosterone also fluctuate across the menstrual cycle, making it difficult to directly attribute these effects to E2 alone. Existing research on brain age in females has primarily focused on hormonal transition phases such as pregnancy and menopause. These phases are characterized by distinct hormonal profiles, and therefore, findings from these populations may not be directly translatable to naturally cycling females. Nevertheless, females who experienced pregnancy (marked by rise in E2 exposure), show less apparent brain aging than nulliparous females (de Lange et al., 2019) But these findings may be influenced by reverse causality, as conception and having children is impacted by maternal pre-pregnancy health and diseases. Also, research in menopausal females indicate that E2 exposure modulates brain age. E2 therapy is linked to lower brain age gap (de Lange et al., 2020) and better cognitive performance (Xu et al., 2024; Zsido et al., 2019). Later onset of menopause (longer E2 exposure) is linked to younger brain age (Luders et al., 2025; Schindler et al., 2022; Subramaniapillai et al., 2022), whereas early menopause, natural or surgical, is associated with structural brain changes (Gervais et al., 2022; Steventon et al., 2023). Yet, despite these largely positive associations between brain age and E2 exposure, the interpretation of E2’s role in naturally cycling females is far from straightforward. Most studies are correlational co-hort studies, which makes a (1) causal interpretation difficult. Additionally, (2) disentangling E2-specific effects from those of other sex hormones that also vary across the menstrual cycle and hormonal status is not possible. Furthermore, findings for association between E2, brain function and aging are not always consistent: while some report beneficial effects of high E2 on cognition (Puri et al., 2025; Andy et al., 2024), others find no associations or even negative outcomes under specific experimental conditions (Gleason et al., 2024; de Lange et al., 2020; Chen et al., 2022). Therefore, systematically assessing the causal impact of E2 is necessary. Menstrual cycle phases with their distinct hormone profiles are linked to dynamic neuroplastic changes across subcortical regions (Pletzer et al., 2019; Denninger et al., preprint MedRx), demonstrating that the female brain is highly plastic and responsive to hormonal modulation. Administration of E2 has been shown to alter brain structure in naturally cycling females and to modulate functional brain networks and activity in emotion and reward processing (Derntl et al., 2024; Matte Bon et al., 2024; Rehbein et al., 2021; Testo et al., 2024; Zsido et al., 2019). For example, E2 administration during the low-hormone phase of the menstrual cycle has been associated with volumetric (Rehbein et al., 2022; Denninger et al., MedRx) and effective connectivity changes (Derntl et al., 2024) in emotion-regulating brain regions, suggesting a direct neuromodulatory role of E2 on affective circuits. In line with this, naturally occurring menstrual cycle-related E2 fluctuations have been associated with changes in emotion recognition (Derntl et al., 2008; Gamsakhurdashvili et al., 2021), emotional competences (Sundström Poromaa & Gingnell, 2014), and emotion regulation—the capacity to modify emotional reactivity (Rehbein et al., 2021). Further, higher E2 levels facilitate the use of adaptive emotion regulation strategies such as reappraisal (Graham et al., 2017), whereas lower E2 levels are associated with increased use of maladaptive emotion regulation strategies such as rumination (Graham et al., 2018). Importantly, rumination and greater worry have also been associated with a higher predicted brain age in late life, whereas other emotion regulation strategies, such as expressive suppression, have shown a protective effect on brain age (Karim et al., 2021). However, it is worth noting that this study included both males and females with and without affective disorders, focusing on individuals aged over 50 years. Additionally, greater psychological resilience has been linked to a more negative BAG in a mixed-sex sample above the age of 53 (Gu et al., 2025) and fluctuating levels of E2 have been also linked to the resilience factor self-esteem (Bloch et al., 1997; Brock et al., 2016; Hill & Durante, 2009; Taylor, 1999). These findings suggest a protective role of resilience factors in brain aging. Given their association with E2 levels, resilient traits may interact with E2 to influence brain age in females.

Taken together, the literature suggests that E2 and traits like the application of beneficial emotion regulation strategies may have a neuroprotective impact against brain aging. However, to the best of our knowledge, a comprehensive study that enables causal interpretations from the effect of E2 on BAG and in association with resilience factors in healthy naturally cycling females is missing so far. Within the current, preregistered study we aimed to investigate the effect of E2 administration on brain aging using a double blinded, randomized crossover placebo-controlled design. We hypothesize that (1) E2 administration will reduce apparent brain age (reflected in a more negative BAG) compared to placebo administration. Due to results indicating an association between brain age and E2 with resilience traits impacting mental health (Karim et al., 2021), we will exploratorily assess whether the association between E2 administration and mental health resilience factors such as self-esteem and emotion regulation traits impact BAG.

## Methods

### Sample

Thirty-two naturally cycling females participated in the study. Menstrual cycle duration of 26 to 32 days was validated by cycle tracking. Exclusion criteria included MRI contraindications (e.g., metal implants), mental, neurological, or endocrine disorders (current or past), hormonal contraceptive use within the last six months, medication intake, or pregnancies. Participants were recruited through the University of Tübingen’s student email system. The Ethics Committee of the Medical Faculty of the University of Tübingen approved the study (754/2017/BO1). All participants provided informed consent and data protection agreements. The study was preregistered (NCT06773429).

The sample size was determined based on power calculations to detect effects of E2 administration on task-based variables (described in Rehbein et al., 2021).

### Procedure

At baseline we collected information on age, depressive symptoms (Becks Depression inventory 2, Beck et al, 1961), anxiety (state and trait anxiety inventory, Spielberg et al., 1970), affect (positive and negative affect schedule (PANAS, Watson et al., 1988), self-esteem (Rosenberg, 1965), and emotion regulation traits (Heidelberg form for emotion regulation strategies (HFERST) & emotion regulation questionnaire (ERQ) (Gross & John, 2003a; Izadpanah et al., 2019). Females were then invited for the first session including neuroimaging (functional magnetic resonance imaging (fMRI)) 2-5 days after menstruation onset. About 24h before the MRI scan, females either orally took a placebo pill or 6 mg of oestradiol valerate (Progynova®21). They took a second dose approximately 6h prior to the MRI scan (see Fig. 1). The E2 dosage was chosen based on a prior study (Bayer et al., 2018), demonstrating that E2 valerate is well-tolerated, safe, and effective in raising E2 levels. The 12-14h half-life of oral E2 valerate allows MRI-scans to be timed when E2 levels peak. In this crossover design, females were invited to the lab again after a minimum of 2 menstrual cycles and switched to the other drug condition (E2 vs. placebo). Blood samples were collected before the first pill intake and immediately before the MRI-scans to measure hormone levels (E2, progesterone (P4), testosterone (T)). Positive and negative affect (PANAS) and state anxiety (STAI) were assessed before the MRI session. See Fig. 1 for the procedure.

**Figure 1.**
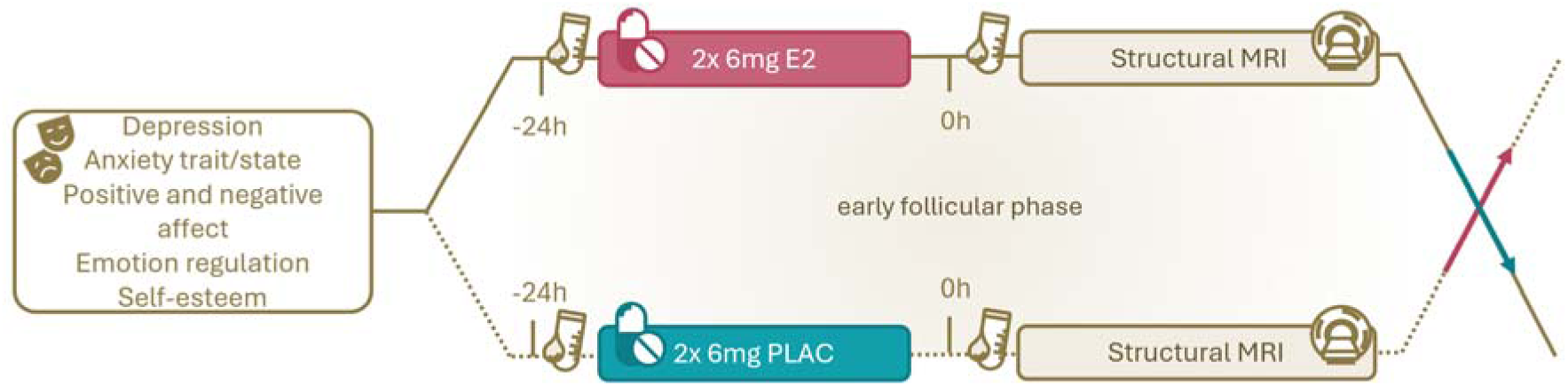
Timeline of the study procedure showing the double-blind, repeated measures crossover design.

### Anatomical MRI data acquisition

MRI data were acquired at a 3 T Siemens PRISMA scanner, University Hospital Tübingen, using a MPRAGE sequence (TR=2300msec, TE=4.16msec, voxel size=1×1×1mm, flip angle=9°, distancing factor 50%, GRAPPA acceleration factor, sagittal orientation, field of view=256×256mm).

### Hormonal analyses

Serum concentrations of E2, P4, T were analysed at the Central Laboratory, University Hospital Tübingen, by use of chemiluminescence assay (CLIA); sensitivity and measurement ranges: E2: 43.60-11,010 pmol/l; P4: 0.67-190.80 nmol/l; T: 0.24-52.05 nmol/l.

### Data analyses

#### MR Data Processing / Quality Control / Brain Age Prediction

We used a pre-trained Simple Fully Convolutional Network brain age model (Leonardsen et al, 2022), available at http://www.github.com/estenhl/pyment-public. This model was previously trained on T1-weighted MRI scans of healthy individuals (total n=53542, 3 to 95 years; female n=27715) from 21 non-overlapping, publicly available datasets. Accordingly, we applied this model to our data, estimating one brain age per scan and visit.

#### Statistical analyses

Data analyses and visualisation were done using MATLAB (R2024a). If normality assumption was violated a Wilcoxon sign-rank test was applied. Hormonal data have previously been reported by Rehbein et al. (2021; 2022) and Derntl et al. (2024). To assess the impact of E2 administration on BAG, we applied linear mixed models (LMMs) with subject as a random effect and age at administration as a covariate-of-no-interest. BAG was calculated as the difference between the chronological age and the predicted age based on the structural brain data. We first tested whether the drug condition (E2 vs. PLAC) had a significant effect on the BAG: BAG ∼ drug administration + age + (1|sub). Then, we tested whether the covariates had an additional impact and included the interaction term drug “administration*covariate” in the LMMs. According to the preregistration, the following covariates were exploratorily assessed (see supplementary): progesterone, testosterone, depression, anxiety, affect, self-esteem, emotion regulation strategies. Analyses were exploratory and hypothesis-generating; accordingly, we report uncorrected p-values and false-discovery-rate corrected p-values (Benjamini & Hochberg, 1995). More information is provided in the supplementary material. To assess the effect of rising E2 levels, the increase in E2 levels (post - pre drug E2 levels, E2Δ) was calculated and applied to LMMs: BAG ∼ E2Δ + age + (1|sub).; BAG ∼ E2Δ*covariates+ age + (1|sub). Finally, the order of drug administration was initially added to the model; however, this did not significantly impact model fit, and to reduce overfitting, order was left out of the final models. If present, outliers were excluded based on the model’s residuals (-3< residuals> 3).

## Results

### Sample Description

28 participants were included in the final analyses. Four females were excluded due to missing data, excessive head movement, or because they started hormonal contraceptives in-between sessions. Days between the two sessions (E2 vs. PLAC condition) ranged from 51-239 days. Please see Table 1 for the sample descriptions.

**Table 1.**
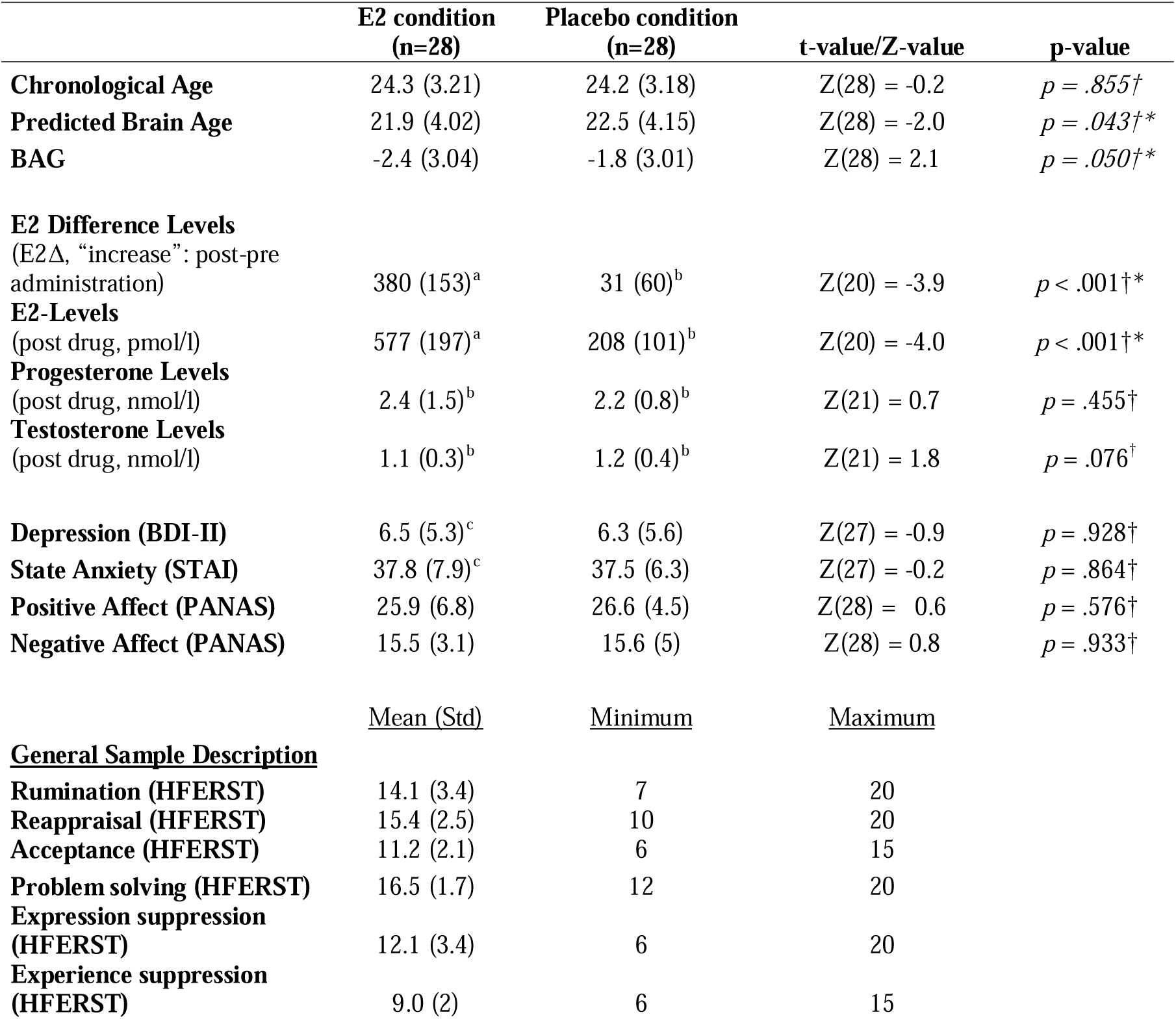
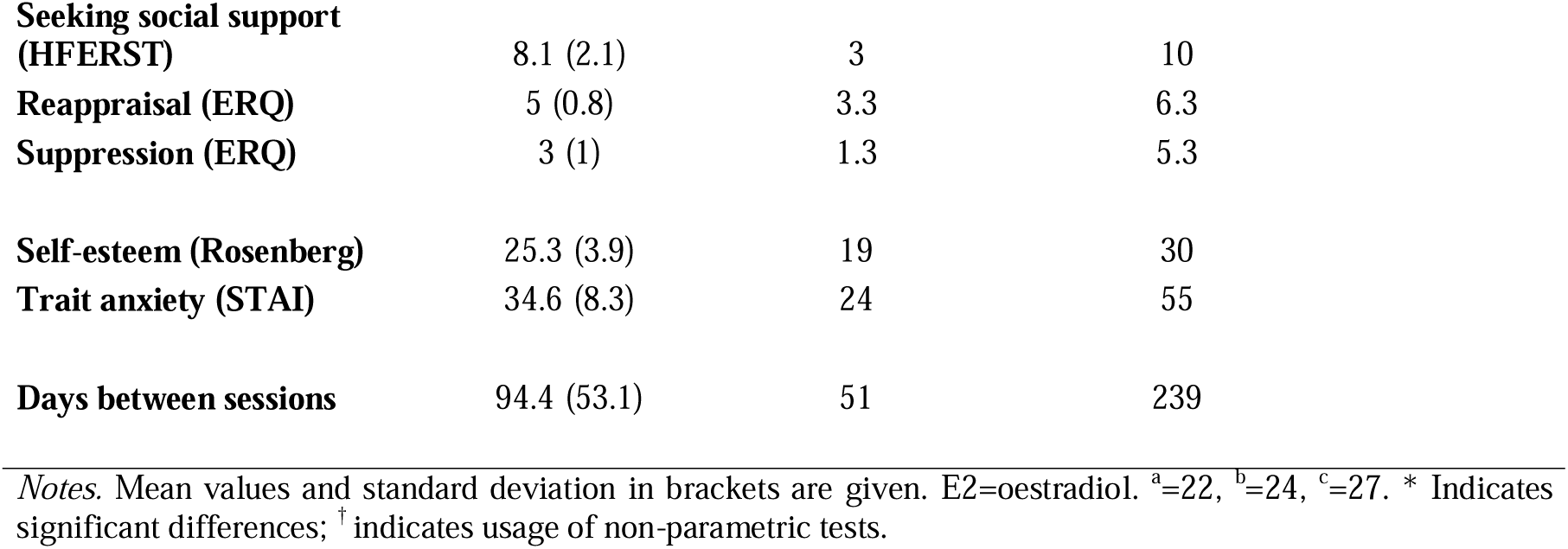
Sample description for both conditions (E2 vs. Placebo).

|  | **E2 condition**<br>**(n=28)** | **Placebo condition**<br>**(n=28)** | **t-value/Z-value** | **p-value** |
| --- | --- | --- | --- | --- |
| **Chronological Age** | 24.3 (3.21) | 24.2 (3.18) | Z(28) = -0.2 | $p = .855^\dagger$ |
| **Predicted Brain Age** | 21.9 (4.02) | 22.5 (4.15) | Z(28) = -2.0 | $p = .043^\dagger*$ |
| **BAG** | -2.4 (3.04) | -1.8 (3.01) | Z(28) = 2.1 | $p = .050^\dagger*$ |
| **E2 Difference Levels**<br>(E2Δ, “increase”: post-pre administration) | 380 (153)<sup>a</sup> | 31 (60)<sup>b</sup> | Z(20) = -3.9 | $p < .001^\dagger*$ |
| **E2-Level**<br>(post drug, pmol/l) | 577 (197)<sup>a</sup> | 208 (101)<sup>b</sup> | Z(20) = -4.0 | $p < .001^\dagger*$ |
| **Progesterone Levels**<br>(post drug, nmol/l) | 2.4 (1.5)<sup>b</sup> | 2.2 (0.8)<sup>b</sup> | Z(21) = 0.7 | $p = .455^\dagger$ |
| **Testosterone Levels**<br>(post drug, nmol/l) | 1.1 (0.3)<sup>b</sup> | 1.2 (0.4)<sup>b</sup> | Z(21) = 1.8 | $p = .076^\dagger$ |
| **Depression (BDI-II)** | 6.5 (5.3)<sup>c</sup> | 6.3 (5.6) | Z(27) = -0.9 | $p = .928^\dagger$ |
| **State Anxiety (STAI)** | 37.8 (7.9)<sup>c</sup> | 37.5 (6.3) | Z(27) = -0.2 | $p = .864^\dagger$ |
| **Positive Affect (PANAS)** | 25.9 (6.8) | 26.6 (4.5) | Z(28) = 0.6 | $p = .576^\dagger$ |
| **Negative Affect (PANAS)** | 15.5 (3.1) | 15.6 (5) | Z(28) = 0.8 | $p = .933^\dagger$ |
|  | <u>Mean (Std)</u> | <u>Minimum</u> | <u>Maximum</u> |  |
| **General Sample Description** |  |  |  |  |
| **Rumination (HFERST)** | 14.1 (3.4) | 7 | 20 |  |
| **Reappraisal (HFERST)** | 15.4 (2.5) | 10 | 20 |  |
| **Acceptance (HFERST)** | 11.2 (2.1) | 6 | 15 |  |
| **Problem solving (HFERST)** | 16.5 (1.7) | 12 | 20 |  |
| **Expression suppression (HFERST)** | 12.1 (3.4) | 6 | 20 |  |
| **Experience suppression (HFERST)** | 9.0 (2) | 6 | 15 |  |

|  |  |  |  |
| --- | --- | --- | --- |
| **Seeking social support (HFERST)** | 8.1 (2.1) | 3 | 10 |
| **Reappraisal (ERQ)** | 5 (0.8) | 3.3 | 6.3 |
| **Suppression (ERQ)** | 3 (1) | 1.3 | 5.3 |
| **Self-esteem (Rosenberg)** | 25.3 (3.9) | 19 | 30 |
| **Trait anxiety (STAI)** | 34.6 (8.3) | 24 | 55 |
| **Days between sessions** | 94.4 (53.1) | 51 | 239 |

*Notes.* Mean values and standard deviation in brackets are given. E2=oestradiol. <sup>a</sup>=22, <sup>b</sup>=24, <sup>c</sup>=27. \* Indicates significant differences; † indicates usage of non-parametric tests.

### Hormonal Data

The increase (E2Δ) as well as the overall E2 levels from pre- to post-administration was significantly higher in the E2- than in the placebo-condition (p>.001). There were no significant differences in progesterone and testosterone levels between the conditions (see Table 1).

### Age differences

Chronological age did not differ between the E2- and PLAC-condition. The predicted age and BAG differed between the conditions, with a younger predicted age and higher BAG at the E2-compared to the PLAC-condition (age: p < .043; BAG: p = .050; Table 1). BAG and chronological age were only weakly correlated (τ = -.075), indicating a weak underestimation of brain age prediction.

### E2 impacts the brain-age-gap

E2 administration and the increase in E2 level were associated with BAG (E2 (reference) vs. placebo: β = 0.60, 95% CI [.04, 1.17], t(53) = 2.13, p = .038, p_FDR_ = .038, η²□ = .079 and E2Δ: β = -.002, 95% CI [-.003, -.0001], t(43) = -2.1433, p = .038, p_FDR_ = .038, η²□ = .097), indicating a significant effect of both E2 administration and E2 increase on the BAG (Figure 2, Table S1 & S2). Nine participants exhibited a larger BAG and nineteen a smaller BAG following E2 compared to placebo administration. Accordingly, exploratory subgroup analyses of these individuals are reported in the supplement (supplementary methods, Figure S2, Table S5-S6).

**Figure 2.**
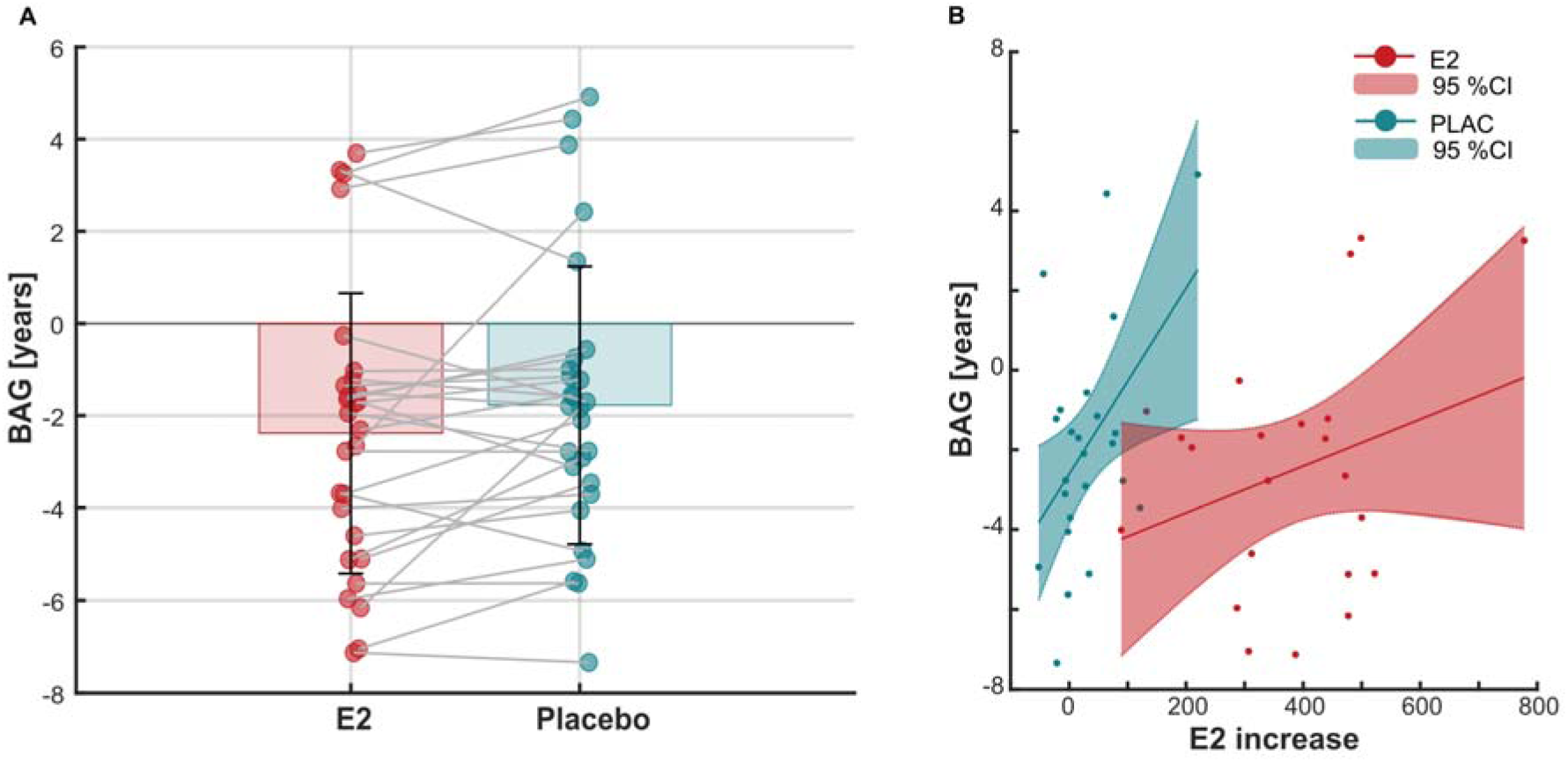
Bar plots showing BAG in years for the E2 (red) and the placebo (blue) drug condition including individual data points and individual trajectories (A). Associations of E2 increase in pmol/l (E2Δ) with BAG in years (B).

### Impact of covariates in association with E2 on brain-age-gap

#### Self-esteem

The model including self-esteem [BAG ∼ drug*self-esteem + age + (1|sub)] showed a significant interaction (β = .15, 95% CI [.01, .29], t(49) = 2.20, p = .033, p_FDR_ = .140, η²□ = .090). However, this effect did not survive correction for multiple comparisons (Figure S1, Tabel S1). Exploratory post-hoc partial correlation analyses did show a negative, although not significant, association between self-esteem and BAG after E2-administration (r(24) = -.114, p = .578) and a positive, but not significant association after placebo administration (r(24) = .073, p = .722). Additionally, the E2 increase and self-esteem interacted on BAG (β = <-.01, 95% CI [<-.01, <-.01], t(39) = -2.18, p = .035, p_FDR_ = .097, η²□ = .109) [BAG ∼ E2Δ*self-esteem + age + (1|sub)] (Table 3), indicating the same pattern. Nevertheless, this interaction likewise failed to remain significant after FDR correction. (Figure S1, Tabel S2).

#### Seeking social support

The model including seeking social support as emotion regulation trait [BAG ∼ drug*social-support + age + (1|sub)] showed a significant main effect of social support (β = -.61, 95% CI [-.13, -.10], t(51) = -2.39, p = .020, p_FDR_ = .225, η²□ = .101) and a significant interaction with drug administration (reference E2: β = .27, 95% CI [.02 .52], t(51) = 2.13, p = .038, p_FDR_ = .140, η²□ = .082). However, both effects did not survive correction for multiple comparisons (Figure S1, Tabel S1). To disentangle the interaction, exploratory post-hoc partial correlation analyses (age as covariate) were conducted which showed a significant negative association between BAG and social support after E2 administration (r(25) = -.424, p = .028), and no effect after placebo-administration (r(25) = -.223, p = .263). Further, social support significantly interacted with E2-increase (β = <-.01, 95% CI [<-.01 <-.01], t(41) = -2.57, p = .014, p_FDR_ = .077, η²□ = .138), showing the same pattern (Figure S1, Tabel S2).

#### Expressive suppression

Expressive suppression of emotions (ERQ) showed a significant main effect of drug administration (reference E2: β = 2.77, 95% CI [1.13 4.41], t(51) = 3.38, p = .01, p_FDR_ = .015, η²□ = .183) and a significant interaction between expressive suppression and drug administration (β = -.71, 95% CI [-1.23 -.20], t(51) = -2.78, p = .008, p_FDR_ = .084, η²□ = .132) (Figure 3, Table S1). Partial correlation analyses did not reveal an association between expressive suppression and BAG during E2 administration (r(25) = .240; p = 228, not significant), or during placebo (r(25) = -.001; p = .996). The interaction effect still indicates that after E2 administration (higher E2-levels) use of expressive suppression is associated with higher brain age, compared to placebo administration (low E2-levels).

**Figure 3.**
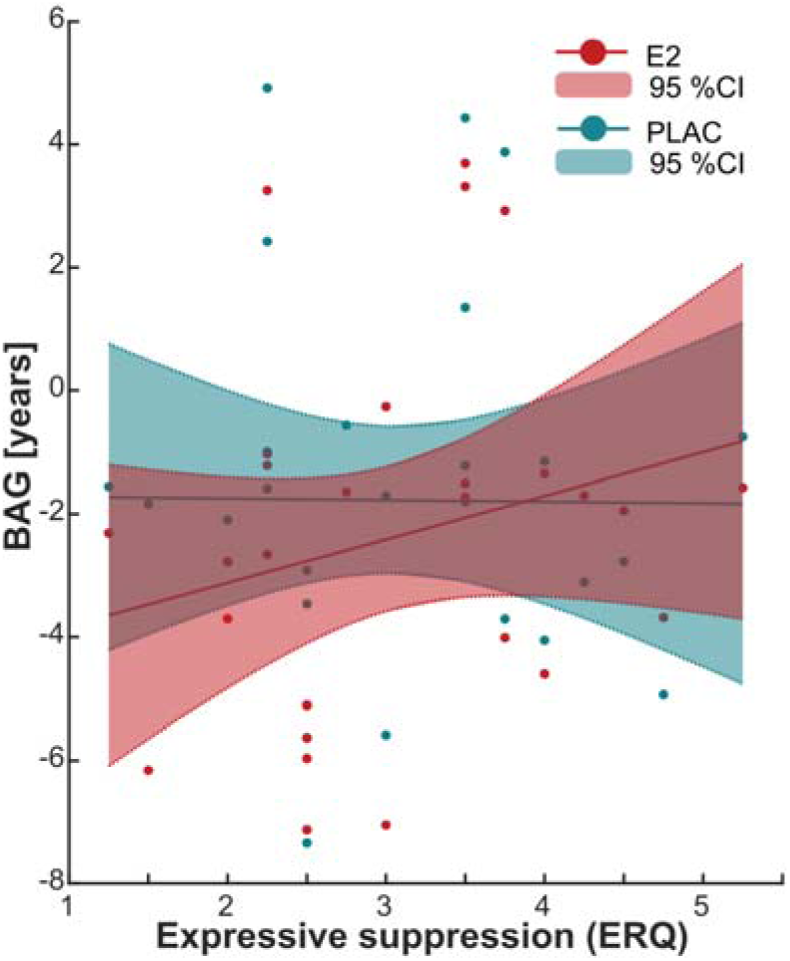
Association expressive suppression measured by the ERQ with BAG in years for E2 (red) and PLAC (blue) drug administration, 95% confident intervals are indicated.

Furthermore, expressive suppression (β = -.01, 95% CI [-.02 -.01], t(41) = -4.35, p = <.001, p_FDR_ = .015, η²□ = .316) showed a main effect of E2 increase and an interaction with E2 increase on BAG (β = <.01, 95% CI [<.01 .01], t(41) = 3.71, p = .001, p_FDR_ = .007, η²□ = .251) (Figure 3, Table S2).

No associations were observed between BAG and covariates we report in our preregistration including emotion regulation, affect, anxiety, depression, and hormone levels (see supplement table S3-S4).

## Discussion

In this study, we analysed the brain age gap (BAG) in relation to oestradiol (E2) levels. To do so, we administered 12 mg of oestradiol valerate to rapidly elevate E2 levels from early follicular to pre-ovulatory levels within 24 hours in a double blind, randomized placebo-controlled crossover design. This study design offers the advantage to assess the effects of E2 on brain aging whilst minimizing variations of other sex hormones that fluctuate across the menstrual cycle (e.g., progesterone). We applied a brain age model that predicts brain age from anatomical MR scans and compared the predicted brain age to the chronological age of each female participant at each time-point. E2 intervention reduced brain age relative to chronological age. Further E2 level increase was associated with the BAG. Additionally, we exploratorily examined how resilience factors may influence the BAG to identify patterns that may guide future studies. Indeed, self-esteem and emotion regulation strategies appear to interact with E2 levels when impacting BAG.

### BAG

Our data shows that the rapid increase of E2 levels within 24 hours after E2 administration leads to a younger appearing brain age compared to lower E2 levels under placebo condition. A higher positive BAG indicates a higher predicted brain age relative to the chronological age. In our data we observed a bias toward negative BAG values, indicating that the brains of naturally cycling females appeared younger than their chronological age in both treatment interventions. However, this effect was stronger after the E2 compared to the placebo administration. The age bias is likely explained by the characteristics of the training dataset from which the prediction model was derived. Since our sample (mean age = 24y) seems younger on average than the training dataset (main contributor UK-Biobank with age range 40-69y), the predicted brain age is underestimated, resulting in systematically lower BAG values.

Our results align well with previous studies. A longer reproductive span, serving as a proxy for cumulative oestrogen exposure, was associated with younger-appearing brains (Luders et al., 2025). Also, the BAG has shown to fluctuate across the menstrual cycle, with brains appearing approximately 1.3 years younger at ovulation (high E2 phase) compared to menses (low E2) and higher E2 levels were correlated with lower predicted brain age (Franke et al., 2015). Although we also observe younger apparent brain age during the E2 intervention, our effect is smaller in magnitude. This variation may arise because we are not achieving the same E2 levels as during the menstrual cycle, which may be partly due to individual differences in hormone metabolism, as orally administered E2-valerate undergoes first-pass metabolism before becoming bioavailable. Our measurements include each treatment intervention across two different menstrual cycles. Although we aimed to minimize variability by administering the intervention during the early follicular phase, hormonal profiles can still differ intra-individually between cycles. Additionally, our analysis focuses on a rapid increase in E2 levels whereas in the menstrual cycle E2 levels may rise more gradually over the mid follicular phase before sharply peaking pre-ovulatory (within 1-2 days) (Schmalenberger et al., 2020). Our data now add to the literature that an E2 administration can rapidly reduce brain age already within 24 hours, suggesting a direct link of E2 and brain age dynamics in naturally cycling females. To derive a brain age prediction, the model derives brain age features directly from structural brain imaging data (including edges, intensity contrasts and tissue characteristics) rather than relying on predefined image features (Leonardsen et al., 2022). These features likely capture aspects of cortical thickness, regional brain volumes, ventricular size, and other structural properties. Features like grey matter volume have shown to be associated with menstrual cycle phase and hormonal levels (Denninger et al., preprint MedRx; Pletzer et al., 2019). Thus, our data now show that E2 affects these brain age features.

The observed effect in the our study might be explained by a rapid E2-dependent synaptogenesis (Khan et al., 2013; Tang et al., 2004) and quick adaptation of oestrogen receptors throughout the brain (Almey et al., 2015; Barth et al., 2015). E2 exerts many of its effects through nuclear oestrogen receptors (ERα and ERβ) as well as membrane-associated receptors (GPER). These receptors are widely expressed throughout the brain, including regions involved in learning and memory formation (e.g., the hippocampus), emotion regulation and motivation (e.g., the amygdala and the basal ganglia), and cognitive control (e.g., the prefrontal cortex) (Barth et al., 2015, 2023). The precise molecular mechanisms by which E2 influences cognitive functions are not yet fully understood. However, E2 has been shown to promote neurotransmitter metabolism, neurotrophin production, and synapse formation, particularly in the hippocampus and prefrontal cortex—regions closely associated with memory, cognition, affect regulation, and stress response (Xu et al., 2024). E2 also affects processes such as cell proliferation, dendritic spine density, synaptic sprouting, axon growth, and myelination. Additionally, it influences major neurotransmitter systems, including the serotonergic, dopaminergic, and GABAergic pathways (for a review, see Barth et al., 2015; 2023). Besides acting via a genomic pathway (over the course of hours to days), E2 can also act rapidly through membrane-bound receptors via a non-genomic pathway, producing effects within minutes (Lai et al., 2017).

### Exploratory effects of resilience factors

Repeated trait use of resilience factors, such as beneficial emotion regulation strategies and self-esteem, can shape brain structure and function through experience-dependent plasticity (Denninger et al., 2025, preprint; Adelstein et al., 2011). The brain’s plasticity has been shown to be adaptive to continued use and training (e.g., in musicians: Krishnan et al., 2013; in sports: Chang et al., 2018). Our explorative data now indicate that this might be the case for resilient factors as well. The trait use of adaptive strategies like high self-esteem and seeking social support may strengthen regulatory neural pathways and thereby promote youth-like brain features, whereas maladaptive strategies such as expressive suppression (Gross & John, 2003a) may contribute to older brain patterns. Our exploratory analysis, suggest that the effect of E2 on BAG is moderated by individual differences in beneficial and maladaptive strategies. Higher self-esteem and social support seem to be nominally linked to a lower BAG (did not survive FDR-correction), whereas expressive suppression is associated with a higher BAG. Although the associations with beneficial strategies did not survive correction for multiple comparisons, the observed pattern aligns well with accounts linking adaptive emotion regulation strategies and psychological resilience to brain aging (Karim et al. 2021; Gu et al 2025). As such, these findings should be interpreted cautiously and viewed as hypothesis-generating rather than confirmatory, warranting replication in larger, adequately powered samples.

#### Beneficial strategies

Both self-esteem and seeking social support represent core resilience factors that promote adaptive coping and mental well-being (Sowislo & Orth, 2013; Schurz et al., 2021). Self-esteem reflects an individual’s sense of self-worth (Rosenberg, 1965), while social support buffers stress and enhances positive affect (Cohen & Wills, 1985; Mulej Bratec et al., 2020; Pei et al., 2023). Our data suggest that both traits interact with endocrinological factors such as E2 administration and E2 increases. Higher self-esteem and greater use of social support seem to be associated with a more negative BAG, indicating younger apparent brain age.

Both self-esteem and social support appear sensitive to E2-related fluctuations (Liparoti et al., 2021; Li & Wang, 2022). For instance, females in the late follicular phase (high E2) show faster approach behaviour toward socio-emotional stimuli compared to the mid-luteal phase, an effect correlated with E2 levels and predictive of social approach motivation (Li & Wang, 2022; Li et al., 2025). Similarly, self-esteem varies across the menstrual cycle, with higher levels observed during the midluteal phase and lower levels around (pre-)menses (Hill & Durante, 2009; Brock et al., 2015).

Self-esteem and social support are known to protect against psychopathology. Low self-esteem is a transdiagnostic risk factor for anxiety and depression (Paxton et al., 2006); conditions linked to accelerated brain aging (Mu et al., 2025). In contrast, higher self-esteem predicts better mental health and may enhance neural resilience through shifts in structural and functional integrity in affective and cognitive control regions, including increased grey matter volume in emotion-regulation areas (Argoskin et al., 2014; Erata et al., 2023). As humans are inherently social, seeking support during stress may similarly protect and shape the resilient brain (Schurz et al., 2021). Our findings align with prior evidence that adaptive emotion-regulation strategies are associated with brain-age alterations (Karim et al., 2021), suggesting that such traits may promote healthier brain age.

Together, these findings point to an interplay between resilience factors and endocrine status that may confer neuroprotective effects. Supporting this view, Gu et al. (2025) demonstrated that higher resilience is associated with a reduced BAG and better cognitive performance in an aged mixed-sex, sample older than 53 years (likely capturing females after menopause), suggesting that resilience factors may have a protective effect on brain age. Extending these findings, our results now indicate that both E2 and resilience factors may contribute to younger brain characteristics.

#### Maladaptive strategies

Expressive suppression refers to the deliberate inhibition of outward emotional expressions despite internal emotional experiences (Gross & John, 2003b). It is generally considered a maladaptive emotion-regulation strategy, consistently linked to heightened negative affect, stress, anxiety, and depressive symptoms, as well as poorer mental health outcomes (Kashdan et al., 2006; Moore et al., 2008; Aldao et al., 2010; Nguyen et al., 2025).

In the present study, greater trait use of expressive suppression of emotions was positively associated with the BAG, suggesting that females who rely less on suppression exhibit a younger apparent brain age. This effect appeared to be further modulated by E2 administration and E2 increase. While little is known about hormonal influences on suppression, emerging evidence indicates that ovarian hormones, particularly E2, can modulate emotion regulation and related neural processes (Graham et al., 2018; Rehbein et al., 2021; Derntl et al., 2024; Denninger et al., MedRx).

In contrast to our findings, Karim et al. (2021) observed a protective role of expressive suppression on brain age in an older, mixed-sex sample, mainly including those with affective disorders. Consequently, female participants were likely menopausal and may have taken hormone replacement therapy, which may have influenced the observed effects. All these factors may shape the impact of expressive suppression on the brain. In contrast, the present study examined young, healthy females with natural menstrual cycles, allowing a direct assessment of hormonal status and emotion regulation traits as potential factors influencing brain age.

#### Limitations and future research

This exploratory, hypotheses-generating study provides novel insights into the effects of exogenous E2 administration on apparent brain aging while minimizing confounding influences of other sex hormones. By focusing on healthy, naturally cycling females, it lays the groundwork for future investigations in broader female populations, including hormonal contraceptive users and clinical groups such as those with premenstrual dysphoric disorder (PMDD). In PMDD, heightened hormonal sensitivity may manifest in brain structural and functional changes (Dubol et al., 2022), thus potentially affecting brain age. E2 administration and resilience factors may present beneficial interventions for these females.

Our study examined short-term E2 increases, whereas most clinical or experimental applications often involve longer-term administration. Future research should determine whether E2-related effects extend to specific cognitive domains such as memory, language, and arithmetic processing. Given E2’s known neurotrophic and neuroprotective properties—promoting hippocampal and basal forebrain cell survival (Morgan et al., 2018), modulating oestrogen-regulated gene transcription (Foster, 2012), and supporting glycolytic metabolism (Vigil et al., 2022)—further work should explore how hormonal context shapes neural aging trajectories and cognitive outcomes.

Notably, orally administered E2 valerate, as used in this study, undergoes first-pass liver metabolism, producing both bioavailable E2 and estrone (Gravelsins & Galea, 2025), with conversion rates depending on formulation and individual metabolism. Different oestrogens—such as E2, ethinyl oestradiol, and estrone—also vary in receptor affinity, metabolism, and downstream effects (Gravelsins & Galea, 2025; Zhu et al., 2006), potentially influencing their impact on brain aging and cognitive function.

Furthermore, the hormone sensitivity hypothesis suggests that some individuals are more responsive to hormonal fluctuations, because of how their neural pathways react to the hormonal shifts (Dubol et al., 2022; Pope et al., 2017). As observed in our study, the exploratory subsample analysis suggests that some females may react more strongly to hormonal shifts than others and may therefore drive more pronounced effects. However, given the limited power due to the small sample size of the subsamples, the interpretability of these findings is constrained. Future research should examine these potential subgroup differences in greater detail.

This differential sensitivity may influence brain age trajectories, highlighting the need for future research to examine how individual variations in hormone responsivity shape brain age trajectories.

Finally, our findings emphasise the contribution of psychological resilience factors, such as self-esteem and emotion regulation, to brain aging. Considering the joint influence of endogenous and exogenous E2 with resilience or risk factors may inform targeted approaches for mental disorders characterized by emotion dysregulation (Kulkarni, 2023; Soares et al., 2001).

## Conclusion

In conclusion, this study provides the first evidence that rapid E2 increases—isolated from other sex hormones—affect estimated brain age. E2 further seems to interact with resilience factors such as self-esteem and social support, with greater use of these adaptive strategies nominally linked to a younger-appearing brain under high levels of E2, whereas reliance on expressive suppression under high levels of E2 corresponded to a higher brain age. These findings underscore the importance of accounting for hormonal dynamics and beneficial as well as maladaptive regulation traits in models of brain age, offering a first step toward integrating neuroendocrine and psychological mechanisms that jointly shape female brain health.

## Data Availability

All data produced in the present study are available upon reasonable request to the authors

## Acknowledgements

We kindly thank Elisa Rehbein for the data collection.

## Author contributions

**AD:** Formal analysis; Data curation; Roles/Writing - original draft & review. **TK:** Methodology; Resources; Software; Validation; Roles/Writing - review & editing. **ER**: Data acquisition; Roles/Writing - review & editing. **ISP:** Conceptualization; Funding acquisition; Project administration; Supervision; Roles/Writing - review & editing. **BD**: Conceptualization; Funding acquisition; Project administration; Resources; Supervision; Roles/Writing - review & editing. **LK**: Conceptualization; Investigation; Data curation; Methodology; Project administration; Supervision; Validation; Visualization; Roles/Writing – original draft, review & editing.

## Declaration of AI use

During the preparation of this work the author(s) used ChatGPT (OpenAI, San Francisco, CA) to assist with language editing. After using this tool/service, the author(s) reviewed and edited the content as needed and take(s) full responsibility for the content of the publication.

